# Neuropsychiatric symptom clusters and plasma proteomic signatures across cognitive ageing and dementia in Kinshasa, Democratic Republic of the Congo: a cross-sectional analysis of the EVCD-RDC cohort

**DOI:** 10.64898/2026.10.26.26366531

**Authors:** Jean Ikanga, Saranya Sundaram Patel, Peng Zhang, Jingjing Yang, Morgen Crosby, Caterina Obenauf, Guy Gikelekele, Emmanuel Epenge, Japhet Magolu Potshi, Topina Tomadia, Francine Manyonga Sabowa, Joseph Phuati Tsangu, Samuel Mampunza, Lelo Mananga, Xuemei Zeng, Alden L. Gross, Alvaro Alonso, Thomas Karikari

## Abstract

**Background:** Neuropsychiatric symptoms (NPS) are common across cognitive ageing and dementia, but their molecular correlates are poorly understood in sub-Saharan African populations. We examined associations between individual NPS, empirically derived NPS dimensions, and plasma proteomic profiles in older adults in Kinshasa, Democratic Republic of the Congo.

**Methods:** In this cross-sectional analysis of the EVCD-RDC cohort, NPS were assessed using the Neuropsychiatric Inventory (NPI) and Geriatric Depression Scale (GDS). Plasma proteins were quantified using the NULISAseq CNS Disease Panel 120. Of 1044 participants in the full cohort, 779 had available proteomic data. Multivariable regression models adjusted for age, sex, education, and estimated glomerular filtration rate, with false-discovery-rate (FDR) correction. Exploratory factor analysis identified latent NPS dimensions.

**Findings:** The full cohort included 1044 participants, of whom 779 comprised the proteomics cohort. Higher NEFL was associated with greater NPI symptom burden (β=0·349, 95% CI 0·201–0·497; FDR<0·001; N=674). Higher IL-6 (β=0·805, 0·362–1·248) and CCL13 (β=0·434, 0·191–0·678) were associated with higher GDS scores (both FDR=0·032; N=693). Higher APOE protein was associated with nighttime behaviour (OR 2·11, 95% CI 1·42–3·11; FDR=0·025). Three NPS dimensions were identified. NEFL was the only factor-level association surviving FDR correction (behavioural/motor: β=0·222, 95% CI 0·127–0·318; FDR<0·001; N=644).

**Interpretation:** NPS showed heterogeneous plasma protein correlates, with the most robust findings implicating neuroaxonal injury and inflammatory signalling. Longitudinal replication in African cohorts is needed.

**Funding:** Alzheimer’s Association; Emory Goizueta Alzheimer’s Disease Research Center; National Institutes of Health.

**RESEARCH IN CONTEXT:** *Evidence before this study:* We searched PubMed and Google Scholar from database inception to Oct 19, 2026, using combinations of the terms “neuropsychiatric symptoms”, “Neuropsychiatric Inventory”, “NPI”, “dementia”, “mild cognitive impairment”, “proteomics”, “protein”, “plasma”, “cerebrospinal fluid”, “cluster”, “subsyndrome”, “Africa”, and “Democratic Republic of the Congo”. We did not restrict the search by language. Existing studies were predominantly observational and conducted in North American or European populations. Most examined overall NPS burden or individual symptoms, particularly depression and apathy. Relatively few studies used high-dimensional proteomic approaches to compare empirically derived NPS dimensions, and evidence from sub-Saharan African populations was largely limited to targeted biomarker studies.

*Added value of this study:* This study extends high-dimensional molecular investigation of NPS to a community-based cohort in the Democratic Republic of the Congo. By examining overall NPS burden, individual symptoms, and empirically derived symptom dimensions alongside multiplex plasma proteomics, we identified robust associations involving NEFL with overall NPS burden and a behavioural/motor dimension, IL-6 and CCL13 with depressive symptoms, and APOE protein with nocturnal behaviour. Most other cluster-level protein associations did not survive correction for multiple testing.

*Implications of all the available evidence:* The molecular correlates of NPS appear heterogeneous and are unlikely to reflect a single neurodegenerative pathway. Blood-based proteomics could help characterise biological processes associated with behavioural manifestations of cognitive ageing and dementia, but replication in independent and longitudinal African cohorts is needed to determine the temporal and clinical significance of these signatures.

## INTRODUCTION

Neuropsychiatric symptoms (NPS) are common and clinically significant manifestations across cognitive ageing, mild cognitive impairment (MCI), and dementia. These symptoms include depression, anxiety, apathy, agitation and aggression, irritability, disinhibition, psychosis, aberrant motor behaviour, sleep disturbance, and appetite changes. Rather than occurring independently, NPS frequently co-occur in recognisable symptom dimensions. Factor-analytic studies have repeatedly identified broad and partially overlapping dimensions involving hyperactivity/agitation, psychosis, affective or mood symptoms, and apathy/neurovegetative symptoms.^1-4^ Although their exact composition varies across populations and statistical approaches, these recurring dimensions provide a useful framework for investigating the clinical and biological heterogeneity of NPS.

The biological mechanisms underlying NPS remain incompletely understood and are unlikely to reflect a single pathological process. Evidence from proteomic and biomarker studies suggests partially overlapping contributions from neurodegenerative, inflammatory, glial, synaptic, and vascular processes.^5-10^ Higher neurofilament light chain (NfL), for example, has been associated with hyperactivity, affective symptoms, and apathy, whereas associations with psychosis and phosphorylated tau biomarkers have been less consistent.^6,7^ These findings suggest that NPS might reflect biological processes extending beyond conventional Alzheimer’s disease pathology alone.

Recent high-dimensional proteomic studies suggest that NPS might also have distinct, although overlapping, molecular signatures.^8-14^ Emerging studies of NPS dimensions have reported differential proteomic and biomarker associations across hyperactivity, affective, apathy, and psychosis domains.^11,12^ Depression and apathy have received comparatively greater study, whereas the molecular correlates of other NPS dimensions remain less well characterised.^5,13,14^ However, relatively few studies have directly used high-dimensional proteomic approaches to characterise and compare empirically derived NPS dimensions. Consequently, whether distinct NPS clusters have reproducible molecular signatures remains unclear.

African populations are also substantially underrepresented in dementia biomarker and proteomic research. Studies from Kinshasa, Democratic Republic of the Congo (DRC), provide initial evidence linking NPS with neurodegeneration-related biomarkers. Higher glial fibrillary acidic protein (GFAP) has been associated with irritability, and GFAP and NfL with depressive symptoms.^15^ A subsequent study similarly identified associations between GFAP, NfL, and geriatric depression.^16^ However, these studies focused largely on targeted biomarkers rather than high-dimensional proteomic profiles. In the present study, we examined associations between individual NPS, empirically derived symptom dimensions from the Neuropsychiatric Inventory (NPI), and plasma proteomic profiles among older adults across cognitive ageing and dementia in Kinshasa. We further evaluated whether distinct NPS dimensions were associated with differential protein signatures, with the aim of characterising the biological heterogeneity of NPS in an understudied sub-Saharan African population.

## MATERIALS AND METHODS

### Study population

Participants were drawn from the EVCD-RDC cohort in Kinshasa, DRC, which has been described previously.^17^ Eligible participants were Congolese citizens residing in the DRC who were aged 65 years or older; provided a home address, telephone number, and contact information for a caregiver or informant; agreed to longitudinal follow-up; were able to provide informed consent independently or through a caregiver; and were fluent in Lingala or French. Individuals with significant visual or auditory impairment or a history of neurodevelopmental, psychiatric, or neurological disorders affecting the central nervous system were excluded. The EVCD-RDC study protocol was approved by the Research Ethics Committee of the School of Public Health at the University of Kinshasa and the Emory University Institutional Review Board. The University of Kinshasa ethics approval was renewed for July 18, 2026, through July 17, 2027; the original approval was issued under protocol ESP/CE/054c/2024. The Emory University protocol was approved under IRB ID MOD006-IRB00113463. All participants provided written informed consent before participation, and study procedures were conducted in accordance with applicable ethical standards and the Declaration of Helsinki

### Clinical and neuropsychiatric assessment

Participants completed the Uniform Data Set version 3, cognitive screening measures, the African Neuropsychology Battery–Short Version, measures of intellectual and functional status, additional questionnaires, and a neurological examination. Instruments were culturally adapted, translated into French and Lingala, and pilot tested before implementation.^17^

NPS were assessed using the Neuropsychiatric Inventory (NPI), an informant-based assessment of 12 symptom domains: delusions, hallucinations, agitation/aggression, depression/dysphoria, anxiety, euphoria/elation, apathy/indifference, disinhibition, irritability/lability, aberrant motor behaviour, nighttime behavioural disturbance, and appetite/eating disturbance.^18^ For the present analyses, symptom presence was coded dichotomously, and the NPI symptom count represented the number of domains present, ranging from zero to 12. Depressive symptoms were assessed using the 15-item Geriatric Depression Scale (GDS-15), with higher scores representing greater depressive symptom burden.^19,20^

### Clinical Adjudication

Participants were classified as cognitively unimpaired (CU), having MCI, suspected Alzheimer’s disease, or other dementia through multidisciplinary consensus involving neuropsychology, neurology, psychiatry, and internal medicine. Diagnostic classification incorporated cognitive performance, functional status, medical history, neurological findings, and clinical course, as previously described.^17,21,22^ Demographically adjusted cognitive scores were based on previously established regression-based reference data from the cohort.^22^

### Plasma Collection and Processing

Blood samples were collected by trained laboratory personnel at the Centre Médical de Kinshasa. Plasma was obtained from EDTA-treated blood following centrifugation at 4000 rpm for 10 min and aliquoted into sterile 0·5 mL tubes. Plasma aliquots were initially stored at –20°C and subsequently transferred to –80°C until biomarker measurement. Additional details of specimen collection and handling have been reported previously.^17^

### NULISA Proteomic Measurement

Plasma proteins were quantified using the NULISAseq CNS Disease Panel 120 (Alamar Biosciences, Fremont, CA, USA) on the ARGO HT platform according to established procedures.^23^ The assay quantified 127 protein biomarkers relevant to neurodegenerative, inflammatory, vascular, metabolic, synaptic, and related biological processes. After thawing, plasma samples were centrifuged at 10 000 g for 10 min and incubated with capture and detection antibodies conjugated to DNA barcodes. Immunocomplexes were purified, corresponding DNA barcodes were ligated, and reporter DNA was quantified by next-generation sequencing. Sequencing counts were normalised to the mCherry internal standard, harmonised using interplate controls, and log₂ transformed to generate NULISA Protein Quantification values. Samples were analysed across ten plates. Vendor-provided sample controls were run in triplicate on each plate, and pooled in-house plasma quality-control samples were run once per plate. Median intraplate and interplate coefficients of variation for the sample controls were 6·3% and 5·8%, respectively; the median interpolated interplate coefficient of variation for the three pooled quality-control samples was approximately 11%. Proteomic analyses included 779 participants with available NULISA data. In the original NULISA quality-control output, 486 samples were classified as Passed and 293 as Warning. Both categories were retained in the analytic dataset based on the QC status provided in the original dataset; no additional sample-level review or exclusion of Warning-status samples was performed.

### Statistical Analysis

Continuous variables are presented as median (IQR), and categorical variables as n/N (%). Characteristics were compared across CU, MCI, suspected Alzheimer’s disease, and other dementia groups using Kruskal–Wallis tests for continuous variables and Pearson χ^2^ tests for categorical variables. The prevalence of each NPS domain was compared across the four diagnostic groups. Pearson χ^2^ tests were used for delusions, agitation/aggression, depression, anxiety, apathy, disinhibition, irritability, nighttime behaviour, and appetite disturbance. Fisher’s exact tests were used for hallucinations, euphoria, and aberrant motor behaviour when expected cell counts were small. These comparisons were descriptive and were not adjusted for multiple testing. For proteomic analyses, separate multivariable linear regression models were fitted for each of the 127 proteins with NPI symptom count and GDS score as continuous outcomes. Individual binary NPS outcomes were analysed using multivariable logistic regression. Models included protein level as the exposure of interest and were adjusted for age, sex, years of education, and estimated glomerular filtration rate (eGFR). Analyses used complete cases for the outcome, protein measure, and covariates; no imputation was performed. The analytic sample was N=674 for the NPI symptom-count models and N=693 for the GDS models. For individual NPS logistic regression models, analytic sample sizes varied by symptom from N=672 to N=680 and were identical across proteins within each symptom. Benjamini–Hochberg false-discovery-rate (FDR) correction was applied separately across the 127 protein tests for each outcome.^24^ FDR-adjusted *p* values below 0·05 were considered statistically significant; nominal associations are reported as exploratory.

### Exploratory Factor Analysis

Exploratory factor analysis was used to identify latent dimensions underlying the 12 binary NPS indicators. Of 1044 participants, 962 had complete symptom data and were included. A Pearson phi correlation matrix was constructed, sampling adequacy was evaluated with the Kaiser-Meyer-Olkin statistic, and Bartlett’s test of sphericity assessed departure from an identity correlation matrix. The number of factors was determined using parallel analysis, comparing observed eigenvalues with the 95th percentiles from 200 simulated binary datasets preserving the observed symptom prevalences. Three factors were retained. Principal-axis extraction with promax oblique rotation was then used, allowing the factors to correlate. Factor labels were assigned descriptively according to the strongest rotated symptom loadings. Factor scores were standardised to mean zero and SD one. Of the 962 participants included in the factor analysis, 720 were also members of the 779-participant proteomics cohort. After restriction to participants with complete covariate data, the final analytic sample was N=644 for each of the three factor-score outcomes. Associations between each factor score and each of the 127 proteins were examined using separate linear regression models adjusted for age, sex, education, and eGFR. FDR correction was applied separately within each factor. Analyses were conducted using R version 4.5.3 (R Foundation for Statistical Computing). Data management and visualisation used tidyverse version 2.0.0 and patchwork version 1.3.2; regression diagnostics used lmtest version 0.9.40 and car version 3.1.5.

## RESULTS

### Participant characteristics and NPS prevalence

The full cohort included 1044 participants: 232 CU, 268 with MCI, 255 with suspected Alzheimer’s disease, and 248 with other dementia; 41 participants without an adjudicated diagnostic status contributed to overall cohort summaries but not diagnostic-group comparisons. Median age was 71·2 years (IQR 67·4–75·7), median education was 10·0 years (6·0–12·0), and 375 of 1038 participants with available sex data were male (36·1%). Age, education, rural residence, MoCA, IDEA-IADL, GDS, and NPI symptom counts differed across diagnostic groups, whereas sex did not (Table 1).

**Table 1.** Characteristics of the study population by neurological status.

| Variable | Overall | CU | MCI | Suspected AD | OD | P value |
| --- | --- | --- | --- | --- | --- | --- |
| **N** | **1044** | **232** | **268** | **255** | **248** | **NA** |
| Age, years | 71.2 (67.4–75.7) | 69.0 (66.0–73.4) | 70.5 (67.4–73.9) | 73.2 (69.0–77.6) | 72.7 (68.7–76.1) | <0.001 |
| Male sex | 375/1038 (36.1%) | 93/232 (40.1%) | 101/267 (37.8%) | 89/254 (35.0%) | 81/248 (32.7%) | 0.348 |
| Education, years | 10.0 (6.0–12.0) | 12.0 (10.0–15.0) | 11.0 (7.0–13.0) | 8.0 (5.0–12.0) | 8.0 (5.0–12.0) | <0.001 |
| Rural residence | 244/1020 (23.9%) | 59/231 (25.5%) | 59/260 (22.7%) | 75/249 (30.1%) | 39/243 (16.0%) | 0.003 |
| MoCA total score | 21.0 (18.0–24.0) | 25.0 (23.0–26.0) | 22.0 (20.0–24.0) | 18.0 (15.0–20.0) | 20.0 (18.0–22.5) | <0.001 |
| IDEA-IADL total score | 28.0 (22.0–32.0) | 30.0 (26.0–33.0) | 29.0 (23.0–33.0) | 25.0 (19.0–29.0) | 28.0 (21.0–32.0) | <0.001 |
| GDS depression score | 4.0 (2.8–6.0) | 3.0 (2.0–5.0) | 4.0 (2.2–6.0) | 4.0 (3.0–7.0) | 5.0 (3.0–7.0) | <0.001 |
| NPI symptom count | 1.0 (0.0–2.0) | 0.0 (0.0–1.0) | 1.0 (0.0–2.0) | 1.0 (0.0–3.0) | 1.0 (0.0–2.0) | <0.001 |

Data are median (IQR) or n/N (%), unless otherwise indicated. *P* values are from Kruskal–Wallis tests for continuous variables and Pearson $\chi^2$ tests for categorical variables; values are unadjusted omnibus comparisons across diagnostic groups. Available-case denominators vary because of missing data. Forty-one participants with missing diagnostic status are included in the Overall column but excluded from diagnostic-group columns and group comparisons. Abbreviations: AD=Alzheimer’s disease; CU=cognitively unimpaired; GDS=Geriatric Depression Scale; IDEA-IADL=Identification and Intervention for Dementia in Elderly Africans–Instrumental Activities of Daily Living; MCI=mild cognitive impairment; MoCA=Montreal Cognitive Assessment; NPI=Neuropsychiatric Inventory; OD=other dementia; NA=not applicable.

Six of the 12 NPS domains differed across diagnostic groups (figure 1). Delusions occurred in 3·9% of CU participants, 5·8% of those with MCI, 10·0% with suspected Alzheimer’s disease, and 5·4% with other dementia (p=0·044). Corresponding proportions were 17·3%, 24·4%, 35·6%, and 31·7% for depression (p<0·001); 2·7%, 3·1%, 12·6%, and 7·4% for apathy (p<0·001); 0·4%, 1·9%, 7·2%, and 2·9% for disinhibition (p<0·001); 15·1%, 19·0%, 29·5%, and 19·8% for irritability (p=0·001); and 11·9%, 22·5%, 39·1%, and 26·3% for appetite disturbance (p<0·001). Participants with suspected Alzheimer’s disease generally had the highest NPS prevalence. Symptom-severity distributions are shown in the appendix. Depression and appetite disturbance had the largest non-zero severity distributions, followed by irritability and agitation/aggression.

**Figure 1.**
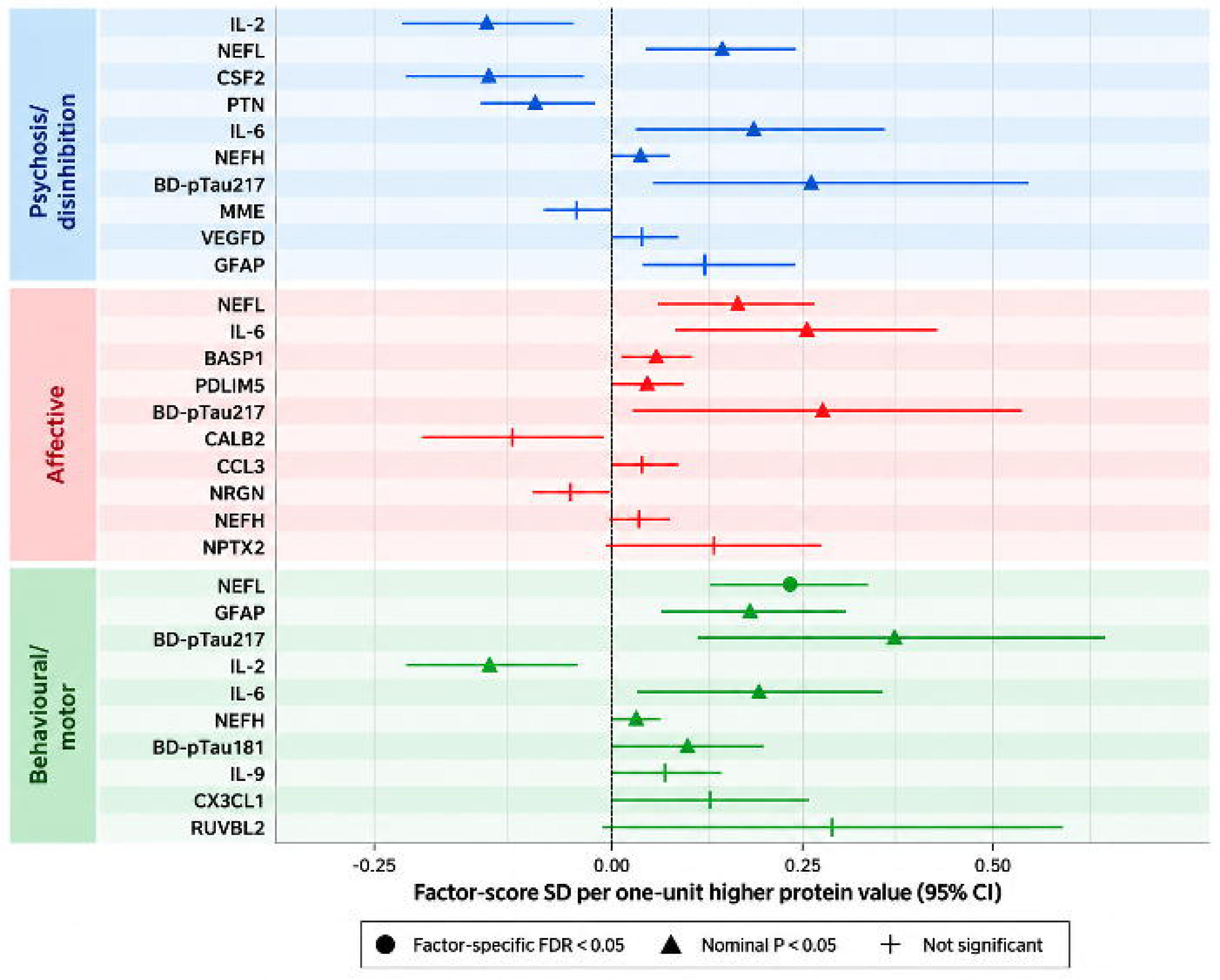
Prevalence of neuropsychiatric symptoms by diagnostic group. Bars show the percentage of participants with each neuropsychiatric symptom among cognitively unimpaired participants (CU), participants with mild cognitive impairment (MCI), suspected Alzheimer’s disease (AD), and other dementia (OD). *p* values represent unadjusted omnibus comparisons across the four diagnostic groups. Pearson χ^2^ tests were used for delusions, agitation/aggression, depression, anxiety, apathy, disinhibition, irritability, nighttime behaviour, and appetite disturbance; Fisher’s exact tests were used for hallucinations, euphoria, and aberrant motor behaviour because of small expected cell counts. Percentages are based on participants with available data for each symptom. Abbreviations: AD= suspected Alzheimer’s disease; CU=cognitively unimpaired; MCI=mild cognitive impairment; NPS=neuropsychiatric symptoms; OD=other dementia.

### Proteomic associations with overall and individual NPS

Of the 1044 participants in the full cohort, 779 had available NULISA proteomic data. After complete-case restriction for outcomes and covariates, 674 participants contributed to the NPI symptom-count models and 693 to the GDS models. After complete-case restriction for outcomes and covariates, 674 participants contributed to the NPI symptom-count models and 693 to the GDS models. Higher NEFL was associated with greater overall NPS burden: each one-unit higher NEFL value was associated with 0·349 additional NPI symptom domains (95% CI 0·201–0·497; FDR<0·001; figure 2A). Higher IL-6 was associated with a 0·805-point higher GDS score (95% CI 0·362–1·248; FDR=0·032), and higher CCL13 with a 0·434-point higher GDS score (0·191–0·678; FDR=0·032). No other proteins remained associated with NPI symptom count or GDS score after FDR correction.

**Figure 2.**
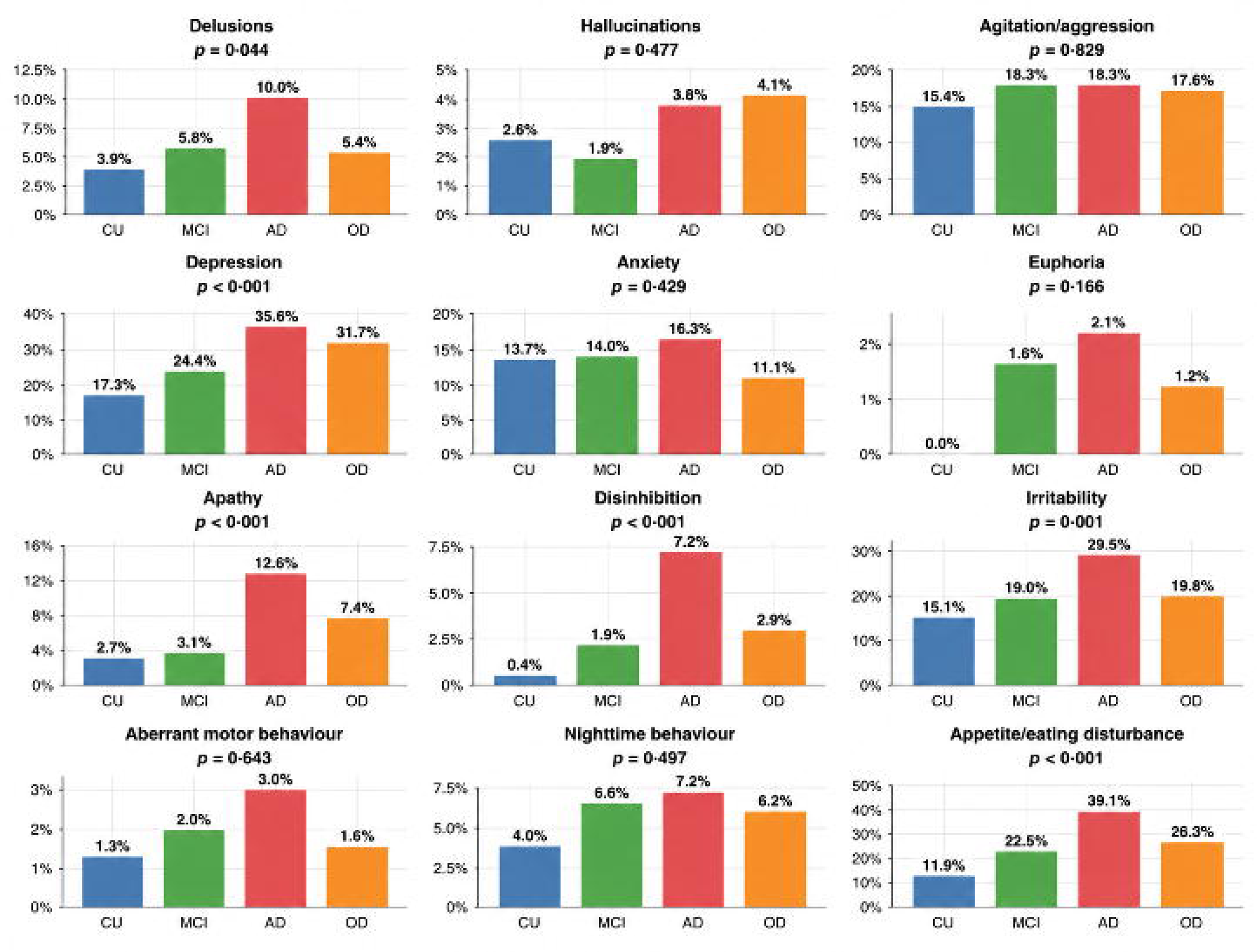
Plasma protein associations with overall and individual neuropsychiatric symptoms. (A) Heatmap of adjusted regression coefficients for the strongest plasma protein associations with Neuropsychiatric Inventory (NPI) symptom count and Geriatric Depression Scale (GDS) score. Positive β coefficients indicate higher NPI symptom count or GDS score with higher protein values; negative coefficients indicate lower scores. The analytic sample was N=674 for the NPI symptom-count models and N=693 for the GDS models. Single asterisks indicate nominal *p*<0·05, and double asterisks indicate associations that remained significant after outcome-specific false-discovery-rate (FDR) correction across 127 proteins. (B) Forest plot of the 20 strongest adjusted plasma protein associations with individual neuropsychiatric symptoms. Estimates are odds ratios (ORs) with 95% CIs for each one-unit higher protein value and are displayed on a logarithmic scale. Individual symptom-model sample sizes ranged from N=672 to N=680. Crosses indicate nominal *p*<0·05, and filled circles indicate associations that remained significant after symptom-specific FDR correction. All models were adjusted for age, sex, years of education, and estimated glomerular filtration rate (eGFR). Abbreviations: CI=confidence interval; eGFR=estimated glomerular filtration rate; FDR=false discovery rate; GDS=Geriatric Depression Scale; NPI=Neuropsychiatric Inventory; NPS=neuropsychiatric symptoms; OR=odds ratio.

Individual symptom models included 672-680 participants, depending on symptom-specific missingness. Among individual NPS, higher APOE protein was associated with nocturnal behaviour (OR 2·11, 95% CI 1·42–3·11; FDR=0·025; figure 2B). No other individual symptom-protein association remained significant after symptom-specific FDR correction. The strongest nominal associations are shown for hypothesis generation in figure 2B and the appendix.

### NPS Factor Structure

Sampling adequacy for factor analysis was acceptable (Kaiser-Meyer-Olkin statistic 0·737), and Bartlett’s test of sphericity indicated that the correlation matrix differed from an identity matrix (χ^2^=846·6, df=66; p<0·001). Parallel analysis supported retention of three factors (figure 3A). The three-factor solution had an RMSR of 0·024 and accounted for 20·6% of total item variance.

**Figure 3.**
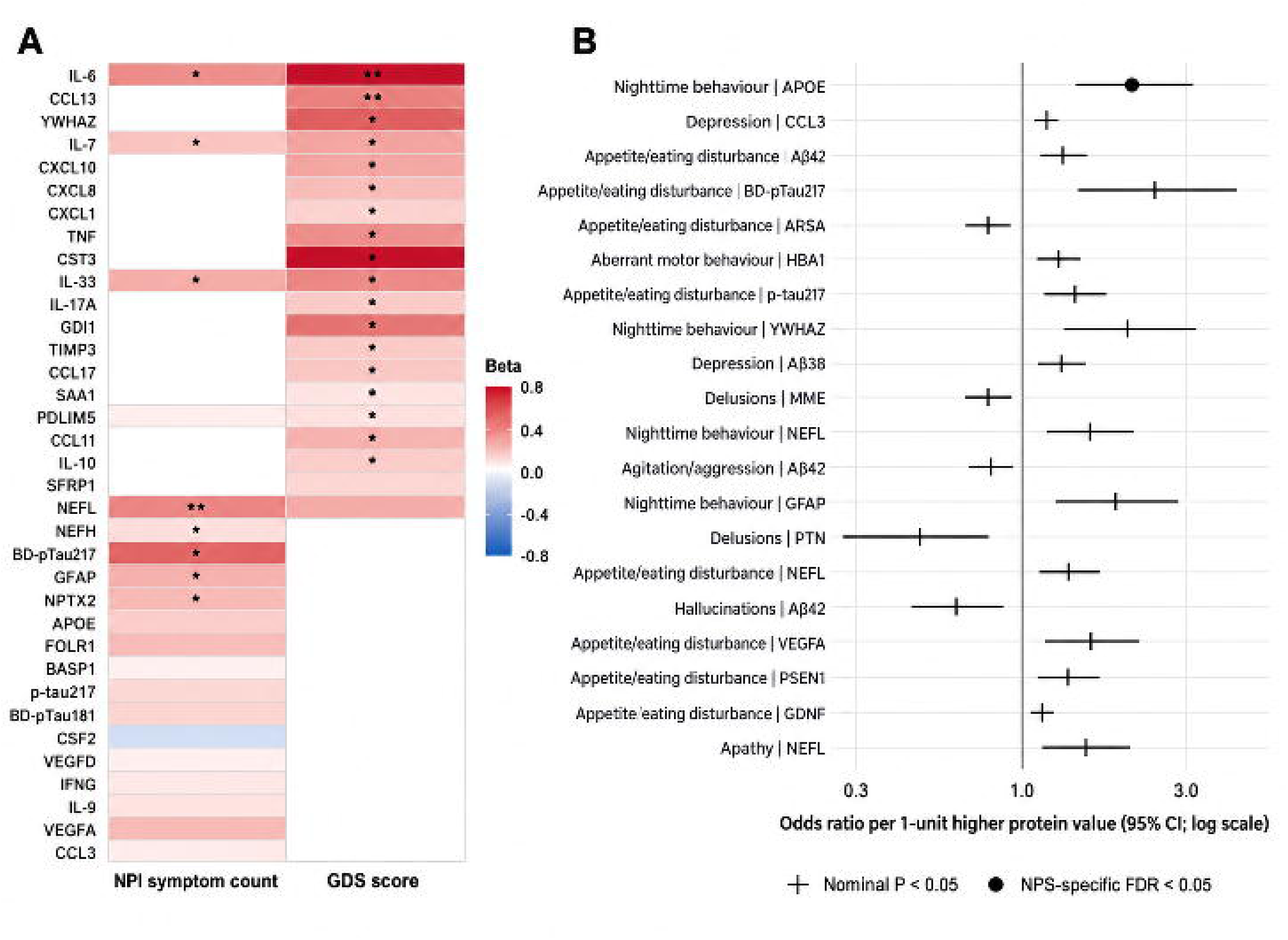
Exploratory factor structure of neuropsychiatric symptoms. (A) Parallel analysis comparing observed eigenvalues with the 95th percentile of eigenvalues from simulated binary datasets. The first three observed eigenvalues exceeded the corresponding simulated thresholds, supporting retention of three factors. (B) Heatmap of promax-rotated factor loadings for the 12 Neuropsychiatric Inventory symptom domains. Larger absolute values indicate stronger associations between individual symptoms and latent factors. Based on the loading patterns, the three factors were labelled psychosis/disinhibition, affective, and behavioural/motor. Abbreviations: NPI=Neuropsychiatric Inventory.

The psychosis/disinhibition factor was characterised most strongly by disinhibition (loading 0·67), delusions (0·55), and hallucinations (0·44). The affective factor was characterised by anxiety (0·46), depression (0·38), and irritability (0·36). The behavioural/motor factor was characterised by nighttime behaviour (0·42) and aberrant motor behaviour (0·34), with appetite disturbance showing a loading of 0·29 (figure 3B). Agitation/aggression, euphoria, and apathy did not load at 0·30 or greater on any factor.

### Proteomic Associations with NPS dimensions

For the psychosis/disinhibition factor, IL-2 and NEFL had the smallest nominal p values (both p=0·005), but neither survived factor-specific FDR correction (both FDR=0·309; figure 4). For the affective factor, the strongest nominal associations were with NEFL (p=0·002; FDR=0·199) and IL-6 (p=0·004; FDR=0·252), neither of which remained significant after correction.

**Figure 4.**
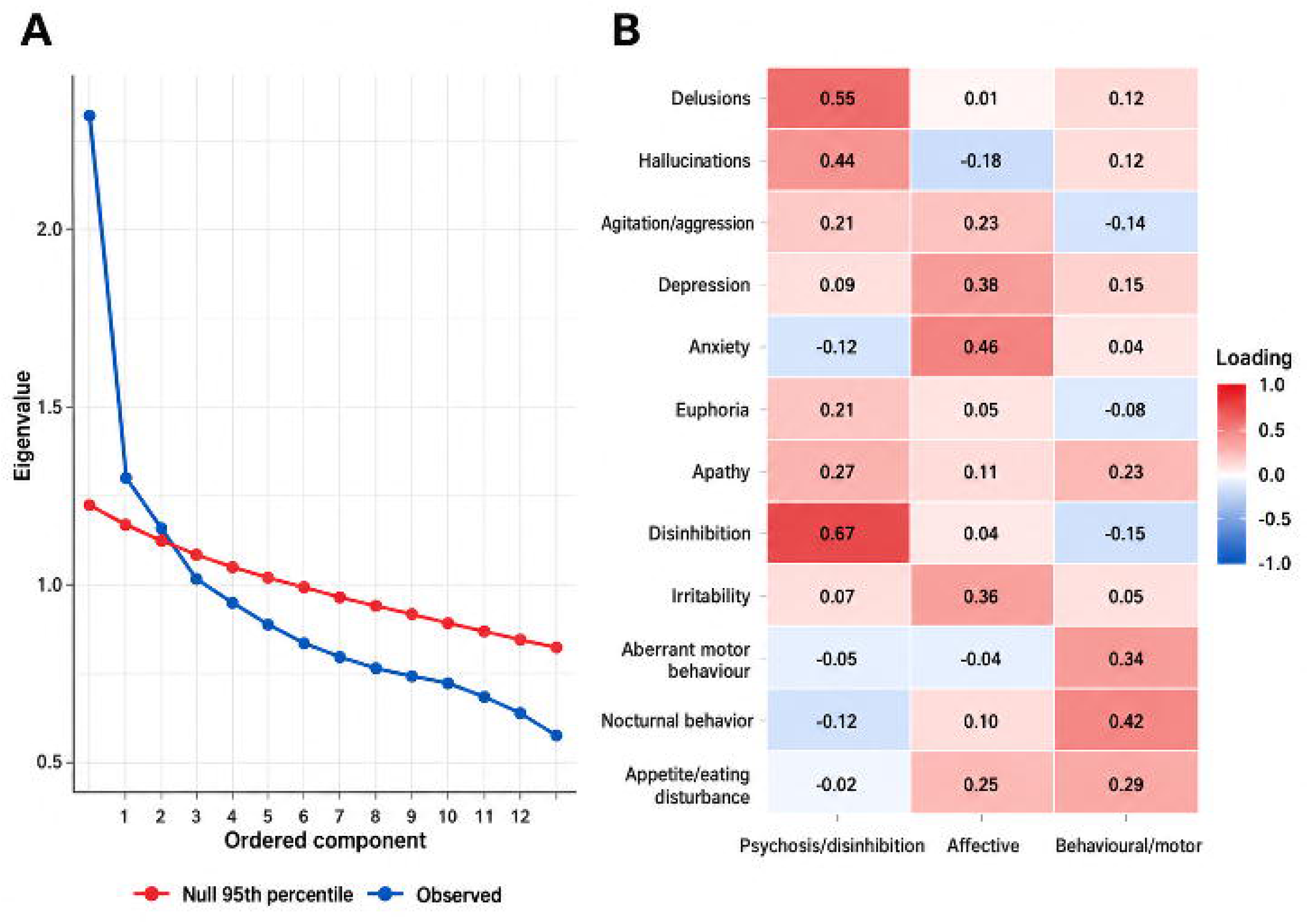
Plasma protein associations with neuropsychiatric symptom factor scores. Forest plot showing adjusted regression coefficients and 95% CIs for the ten strongest plasma protein associations with each neuropsychiatric symptom factor score. Factor scores were standardised to mean 0 and SD 1. Estimates represent the SD difference in factor score associated with each one-unit higher protein value. Protein-factor models included N=644 participants for each of the three factor-score outcomes. Models were adjusted for age, sex, years of education, and estimated glomerular filtration rate (eGFR). Filled circles indicate associations that remained significant after factor-specific false-discovery-rate (FDR) correction across 127 proteins; triangles indicate nominal *p*<0·05; and plus signs indicate displayed associations that were not nominally significant. The vertical line at zero indicates no association. Abbreviations: CI=confidence interval; eGFR=estimated glomerular filtration rate; FDR=false discovery rate; NPS=neuropsychiatric symptoms; SD=standard deviation.

Among 644 participants with complete proteomic, factor-score, and covariate data, higher NEFL was associated with a higher behavioural/motor factor score (β=0·222, 95% CI 0·127–0·318; p<0·001; FDR<0·001) and was the only factor-level protein association that remained significant after FDR correction.

## DISCUSSION

In this cross-sectional study of older adults in Kinshasa, plasma proteomic profiles were associated with overall NPS burden, depressive symptoms, selected individual NPS, and empirically derived behavioural dimensions. The most robust findings after correction for multiple testing involved higher NEFL with overall NPS burden and the behavioural/motor factor, higher IL-6 and CCL13 with depressive symptoms, and higher APOE protein with nighttime behaviour. Most other symptom-specific and factor-level associations were nominal, underscoring the biological heterogeneity of NPS and the need for independent replication.

The three empirically derived NPS dimensions—psychosis/disinhibition, affective, and behavioural/motor—partly overlapped with dimensions identified in previous factor-analytic studies.^1-4^ However, several symptoms did not load clearly on any factor, and the three-factor solution explained only 20·6% of total item variance. These findings are consistent with previous evidence that broad NPS dimensions recur across cohorts but that the placement and strength of individual symptoms are not fully stable. This variability might reflect differences in clinical characteristics, symptom prevalence, sociocultural context, or statistical approaches and cautions against assuming that similarly labelled NPS clusters represent identical constructs across populations.

The strongest proteomic findings implicated neuroaxonal injury and inflammatory signalling. NEFL was associated both with greater overall NPS burden and with the behavioural/motor dimension, consistent with previous reports linking NfL with several behavioural manifestations of cognitive impairment.^6,7^ IL-6 and CCL13 were associated with depressive symptom burden, supporting a possible contribution of inflammatory and immune processes to affective symptoms. Higher APOE protein was associated with nighttime behaviour, although the biological significance of this association requires replication. In contrast, the broader patterns involving amyloid/tau, vascular, metabolic, glial, and synaptic proteins were predominantly nominal and should be viewed as hypothesis-generating rather than evidence of established cluster-specific pathways.

These findings extend previous biomarker work in the DRC, in which GFAP and NfL were associated with selected NPS and depressive symptoms.^15,16^ Interpretation of circulating inflammatory and neurodegenerative proteins in this setting nevertheless requires particular caution. Infectious, nutritional, vascular, metabolic, and environmental exposures might influence systemic immune signalling and plasma protein concentrations independently of central nervous system pathology. Peripheral inflammatory signals therefore should not be equated with neuroinflammation without complementary central biomarkers, imaging, or neuropathological evidence. At the same time, blood-based multiplex proteomics offers a minimally invasive approach for studying multiple biological systems in settings where cerebrospinal fluid sampling and advanced neuroimaging can be difficult to implement.

This study has several strengths, including detailed neuropsychiatric phenotyping, examination of both individual symptoms and empirically derived dimensions, multiplex measurement of 127 plasma proteins, and inclusion of a large and substantially understudied sub-Saharan African cohort. Several limitations should also be considered. First, the cross-sectional design precludes conclusions about temporal or causal relationships. Second, although FDR correction reduced false-positive findings, the large number of proteins and NPS outcomes means that nominal associations require replication. Third, preanalytical factors, including sample collection, storage, transport, and haemolysis, can influence circulating protein measurements. Haemolysis might be particularly important in settings where infections associated with haemolysis, including malaria, are prevalent. Fourth, population-specific reference data for circulating proteomic measures in older adults in sub-Saharan Africa remain limited, and differences from cohorts studied elsewhere could reflect environmental, infectious, genetic, physiological, or structural factors rather than disease-specific processes. Finally, the NPI has not been comprehensively validated in this population or across relevant local linguistic and cultural contexts.

In conclusion, NPS in cognitive ageing and dementia were associated with heterogeneous plasma protein signatures in this cohort from Kinshasa. The most robust findings implicated neuroaxonal injury, inflammatory signalling, and APOE-related biology, whereas most cluster-specific associations were exploratory. Longitudinal and independent replication in African populations will be important to establish whether these signatures precede, accompany, or result from NPS and whether they contribute clinically useful information beyond cognitive diagnosis.

## Data Availability

All data produced in the present study are available upon reasonable request to the authors

## CONTRIBUTORS

JI conceived and supervised the study and contributed to study design, participant recruitment, clinical assessment, interpretation of the findings, and manuscript preparation. SSP contributed to the literature review, interpretation of findings, and drafting and revision of the manuscript. PZ and JY conducted the statistical analyses and contributed to interpretation of the results and revision of the manuscript. MC contributed to the literature review and manuscript revision. CO contributed to study coordination and manuscript revision. GG, EE, JMP, TT, FMS, JPT, SM, and LM contributed to participant recruitment, clinical assessment, cohort implementation, and interpretation of the findings. XZ and TK contributed to proteomic measurement, quality control, and interpretation of proteomic findings. ALG contributed to funding acquisition and critical revision of the manuscript. AA contributed to study interpretation and critical revision of the manuscript. PZ and JY directly accessed and verified the underlying participant-level data. All authors had access to the study data relevant to their contributions, reviewed and approved the final manuscript, and accept responsibility for the decision to submit the manuscript for publication.

## DATA SHARING

Deidentified participant-level data underlying the findings reported in this Article, together with a data dictionary, may be made available to qualified researchers following publication upon reasonable request to the corresponding author and approval by the EVCD-RDC study investigators and relevant ethics committees. Requests should include a research proposal describing the intended analyses. Data sharing will be subject to applicable participant-consent, institutional, ethical, and data-use requirements.

## ACKNOWLEDGMENTS

We thank the participants, families, caregivers, community partners, clinical staff, and research personnel in Kinshasa whose participation and contributions made this study possible.

