## Supplementary Appendix for "Neuropsychiatric symptom clusters and plasma proteomic signatures across cognitive ageing and dementia in Kinshasa, Democratic Republic of the Congo: a cross-sectional analysis of the EVCD-RDC cohort"

Contents: Supplementary Figure 1 - Severity distribution of neuropsychiatric symptoms (p 1); Supplementary Figure 2 - Plasma protein associations across individual neuropsychiatric symptoms (p 2).

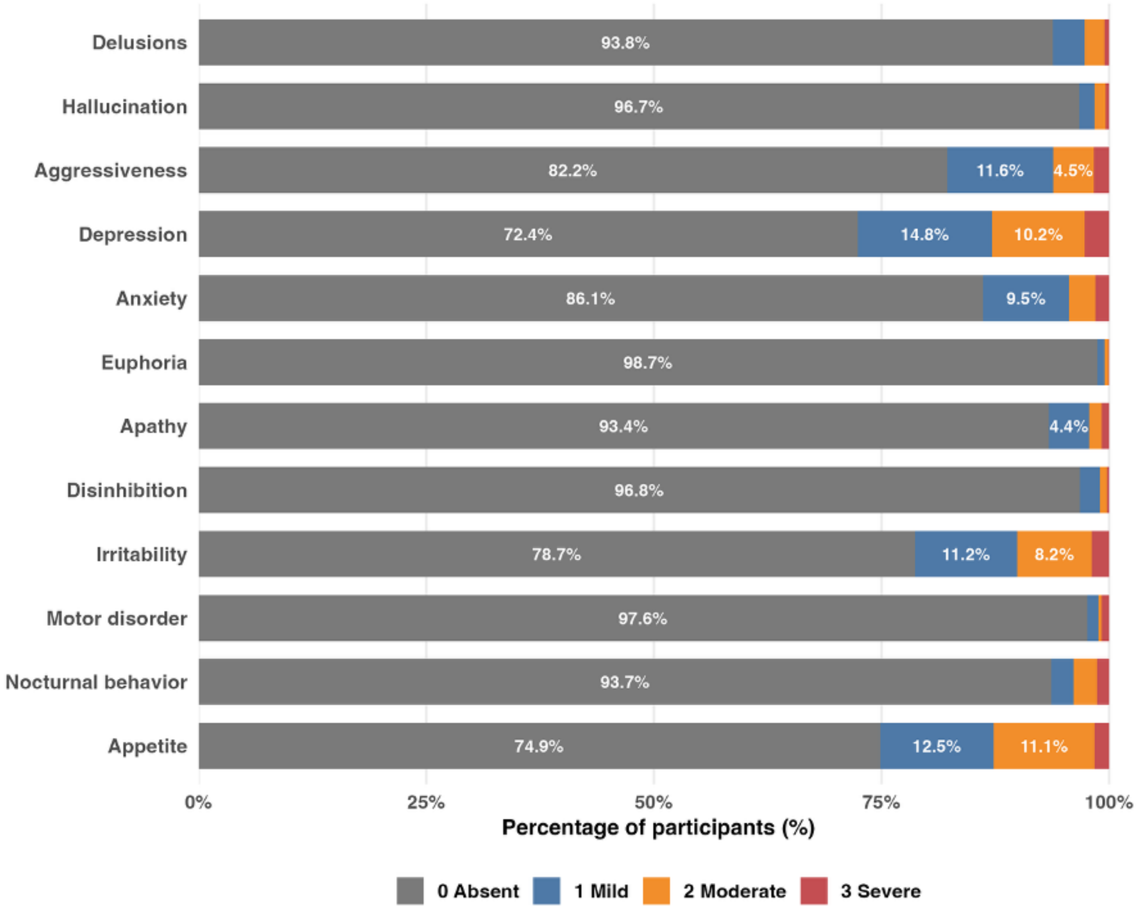


**Supplementary Figure 1. Severity distribution of neuropsychiatric symptoms.**
Stacked bars show the distribution of symptom severity among participants with observed data for each neuropsychiatric symptom. Symptoms were coded as 0=absent, 1=mild, 2=moderate, and 3=severe. Percentages are calculated among participants with non-missing symptom data; missing observations are not included in the absent category.

### Supplementary appendix

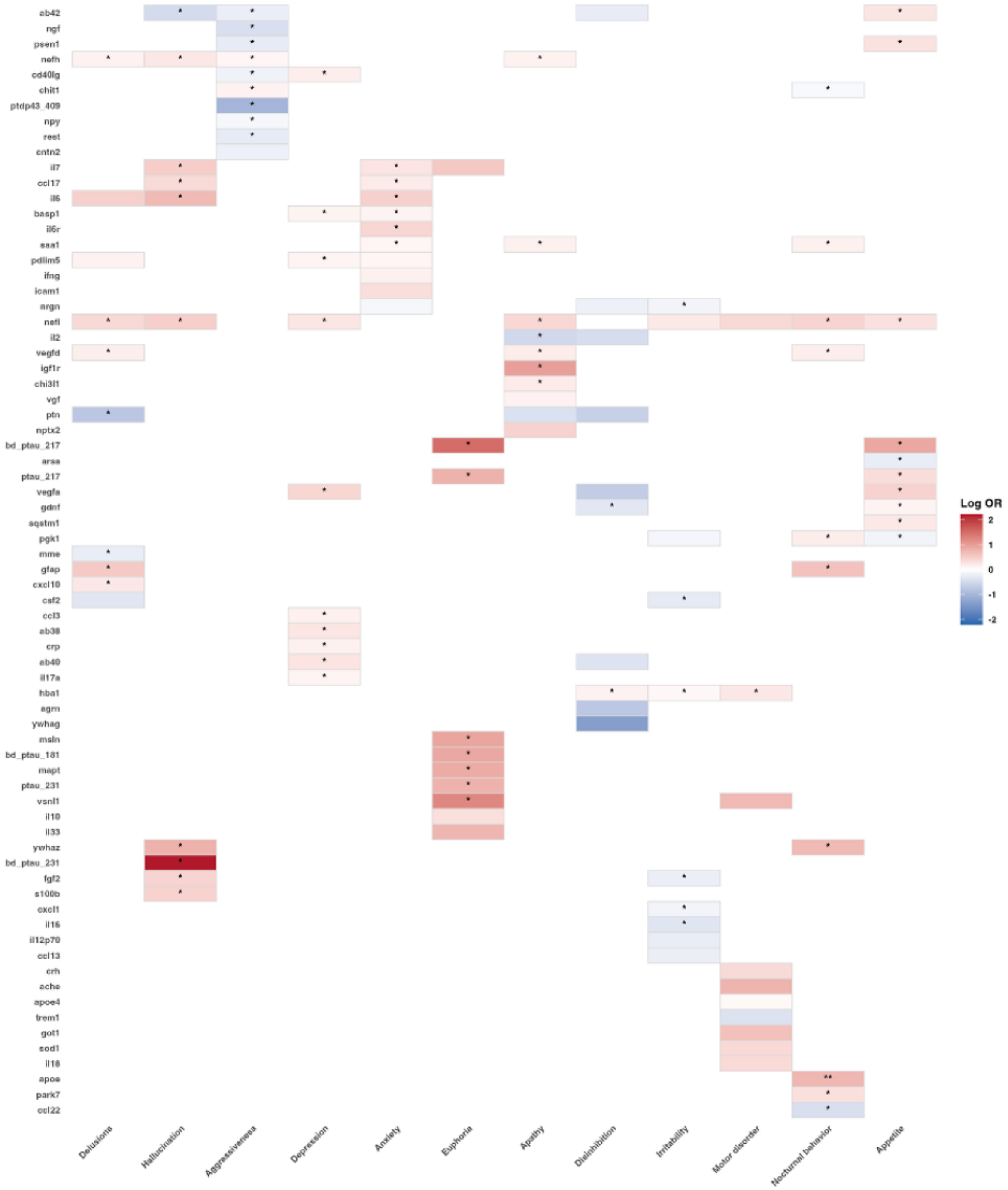


**Supplementary Figure 2. Plasma protein associations across individual neuropsychiatric symptoms.**
Heatmap showing adjusted log odds ratios for associations between plasma proteins and individual neuropsychiatric symptoms. Models were adjusted for age, sex, years of education, and estimated glomerular filtration rate (eGFR). Positive log odds ratios indicate higher odds of the corresponding symptom with higher protein values, whereas negative values indicate lower odds. Single asterisks indicate nominal p<0·05; double asterisks indicate associations that remained significant after symptom-specific false-discovery-rate (FDR) correction across 127 proteins. Blank cells indicate associations not selected for display.
